# Shared Genetic Architecture Between Parkinson’s Disease and Sleep-Related Traits Implicates the *MAPT* Locus on Chromosome 17

**DOI:** 10.1101/2025.06.19.25329954

**Authors:** Aura Aguilar-Roldán, Miguel E. Rentería, Luis M. García-Marín

## Abstract

Parkinson’s disease (PD) is a neurodegenerative disorder characterised by both motor and non-motor symptoms. Among the latter, sleep disturbances are particularly common and include insomnia, obstructive sleep apnoea, excessive daytime sleepiness, restless legs syndrome, and REM sleep behaviour disorder. In this study, we investigated the shared genetic architecture between PD and sleep-related traits to uncover biological pathways that may underpin this relationship. We analysed genome-wide association study (GWAS) summary statistics for PD (∼31,700 cases, ∼18,600 proxy cases, ∼1.4 million controls) and eight self-reported sleep-related traits (each with n > 300,000): ease of getting up, chronotype (morningness), napping, insomnia, obstructive sleep apnoea, snoring, daytime dozing, and sleep duration. Genetic correlations were estimated using LD score regression, and GWAS-Pairwise analysis was used to identify genomic segments harbouring shared causal variants. We then mapped these variants to protein-coding genes using MAGMA. We observed a significant genome-wide genetic correlation between PD and daytime dozing (P < 0.05). At the local level, six genomic regions contained shared variants. A single locus on chromosome 17 contributed the majority of mapped protein-coding genes, including *ARHGAP27*, *PLEKHM1*, *CRHR1*, and *MAPT*, which are implicated in neurodegeneration and circadian rhythm regulation. These findings suggest that the *MAPT* locus, beyond its established role in PD, may also contribute to sleep-wake regulation via shared biological pathways, including tau pathology, stress response, and chromatin remodelling. Our results highlight sleep disturbances as a potential early marker of, or risk factor for, Parkinson’s disease susceptibility.

## INTRODUCTION

Parkinson’s disease (PD) is a progressive neurodegenerative disorder that affects approximately 2% of individuals over 60 years old^1^. While PD is primarily characterised by motor symptoms such as bradykinesia, tremor, and rigidity, it also manifests through a broad range of non-motor symptoms, including cognitive impairment, mood disorders, and sleep disturbances. The disease is strongly associated with the progressive degeneration of dopaminergic neurons in the substantia nigra, driven by the pathological accumulation of alpha-synuclein aggregates^2^. Notably, these pathological processes have also been implicated in regulating sleep-wake cycles^3^.

Sleep disturbances are among the most prevalent non-motor symptoms in PD, affecting between 60% and 98% of individuals with the disorder^4,5^. Common sleep-related symptoms in PD include insomnia, excessive daytime sleepiness, restless legs syndrome, nocturnal akinesia, rapid eye movement sleep behaviour disorder (RBD), circadian rhythm disruptions, and obstructive sleep apnoea (OSA)^6,7^. The prodromal presence of RBD has been associated with a more severe motor and non-motor PD subtype, suggesting a potential disease-modifying role of this parasomnia^8^. A large prospective, population-based study demonstrated that subjectively poor sleep quality and shorter sleep duration were associated with an increased risk of incident Parkinsonism within the first two years of follow-up^9^. These findings underscore the importance of investigating sleep disturbances and their potential shared genetic basis with PD, as they may serve as early biomarkers of disease onset, particularly in individuals who are genetically susceptible.

Genetic factors are known to contribute to individual differences in the aetiology of both PD and sleep-related traits. Twin and family studies have reported moderate to high heritability estimates for sleep phenotypes, including sleep duration (0.46^10^, insomnia (0.38–0.59^11^), and napping 0.65^12^). Likewise, the heritability for PD has been estimated at 0.34^13^. Genome-wide association studies (GWAS) have identified more than 90 genetic risk loci associated with PD^14^, and similarly large numbers of loci have been linked to sleep-related traits^15^, with one GWAS for insomnia reporting up to 554 risk loci^16^.

Although emerging evidence suggests that the interplay between neurodegeneration, neurotransmitter imbalances, and genetic factors may underlie the connection between PD and sleep, the genetic architecture of this relationship remains poorly understood. In this study, we leveraged GWAS summary statistics for PD and eight sleep-related traits (OSA, daytime dozing, ease of getting up, sleep duration, napping, insomnia, morningness, and snoring) to identify shared genetic loci and map associated variants to protein-coding genes. Through this approach, we aimed to elucidate the genetic basis of the relationship between sleep and PD, providing new insights into the underlying biological pathways and contributing to efforts toward early detection and intervention.

## METHODS

### GWAS Summary Statistics for Parkinson’s Disease

We analysed GWAS summary statistics for PD in individuals of European ancestry. These data were derived from the International Parkinson’s Disease Genomic Consortium (IPDGC), 23andMe, and the UK Biobank. The dataset comprised approximately 31,700 PD cases, 18,600 proxy cases from the UK Biobank (i.e., individuals with a first-degree relative diagnosed with PD), and 1,417,791 controls, totalling 1,474,097 participants. A meta-analysis was performed using a fixed-effects model implemented in METAL^17^. Access to the 23andMe cohort was obtained through the appropriate application process (https://research.23andme.com/dataset-access/) and governed by an institutional data transfer agreement. Further details regarding GWAS design and quality control procedures can be found in the original publication ^14^.

### GWAS Summary Statistics for Sleep-Related Traits

We used GWAS summary statistics from individuals of European ancestry for eight self-reported sleep-related traits: obstructive sleep apnoea (UK Biobank, *n*=523,366), snoring (UK Biobank, *n*=408,317), insomnia (UK Biobank and 23andMe, *n*=2,365,010), daytime dozing (UK Biobank, *n*=386,548), ease of getting up (UK Biobank, *n*=385,949), sleep duration (UK Biobank, *n*=384,317), napping (UK Biobank, *n*=386,577), and morningness (UK Biobank, *n*=345,552). Participants with missing or ambiguous responses (e.g., "prefer not to answer" or "I don’t know") were excluded. Summary statistics were processed using strict quality control criteria for imputation quality and for minor allele frequency (MAF). Full details of the cohorts and quality control procedures are available in the original publications^15,18,19^.

### Genetic Correlations

We estimated genome-wide genetic correlations between PD and each of the eight sleep-related traits using linkage disequilibrium score regression (LDSC)^20^. This approach quantifies the extent to which genetic effects are shared across traits by correlating GWAS effect sizes across the genome. To account for multiple comparisons, we applied a Bonferroni correction, setting the significance threshold at p < 0.05/8 (number of sleep-related traits tested).

### Pairwise Analysis of GWAS

To identify genomic regions harbouring variants that influence both PD and at least one sleep-related trait, we conducted pairwise GWAS (GWAS-PW) analysis. This method divides the genome into ∼1,700 independent regions based on linkage disequilibrium patterns and calculates the posterior probabilities of four models: (i) association with PD only; (ii) association with a sleep-related trait only; (iii) shared association with both PD and a sleep-related trait via common causal variants; and (iv) distinct causal variants for each trait ^21^. Genomic segments with a posterior probability of association (PPA) > 0.5^22–25^ under model (iii) were prioritised for downstream analyses, in line with prior work.

### Multivariate Analysis of Genomic Annotation

We used MAGMA v1.08^26^ within the Functional Mapping and Annotation (FUMA) platform v1.6.0^27^ to further investigate the genomic segments identified through GWAS-PW. Specifically, we conducted gene-based and gene-set analyses to map variants in these regions to protein-coding genes jointly associated with PD and sleep-related traits. For the PD gene-set analysis, we applied a Bonferroni correction based on the total number of gene sets tested (N = 17,008). For napping and ease of getting up, gene-set significance was corrected for the number of genes in the shared genomic segments^28^.

### Protein–Protein Interaction (PPI) Network Analysis

We performed PPI network analysis using the STRING database v12.0^29^ to examine biological interactions in the most relevant shared loci. We included significant protein-coding genes mapped by MAGMA and retained medium and high-confidence interactions with a score ≥ 0.4. We also applied the Markov Cluster Algorithm (MCL) with an inflation parameter of 3 to identify potential functional modules. Network enrichment was assessed by comparing the observed number of edges with the expected number given the size of the input set, using STRING’s built-in enrichment test.

## RESULTS

We estimated genome-wide genetic correlations between sleep-related traits and Parkinson’s disease (PD) using linkage disequilibrium score regression. Daytime dozing showed a nominally significant negative genetic correlation with PD (rG = -0.1, P-value = 0.0485). However, after correcting for multiple testing, no genetic correlations between PD and the sleep-related traits reached statistical significance (Figure 1; Supplementary Table 1).

**Fig 1.**
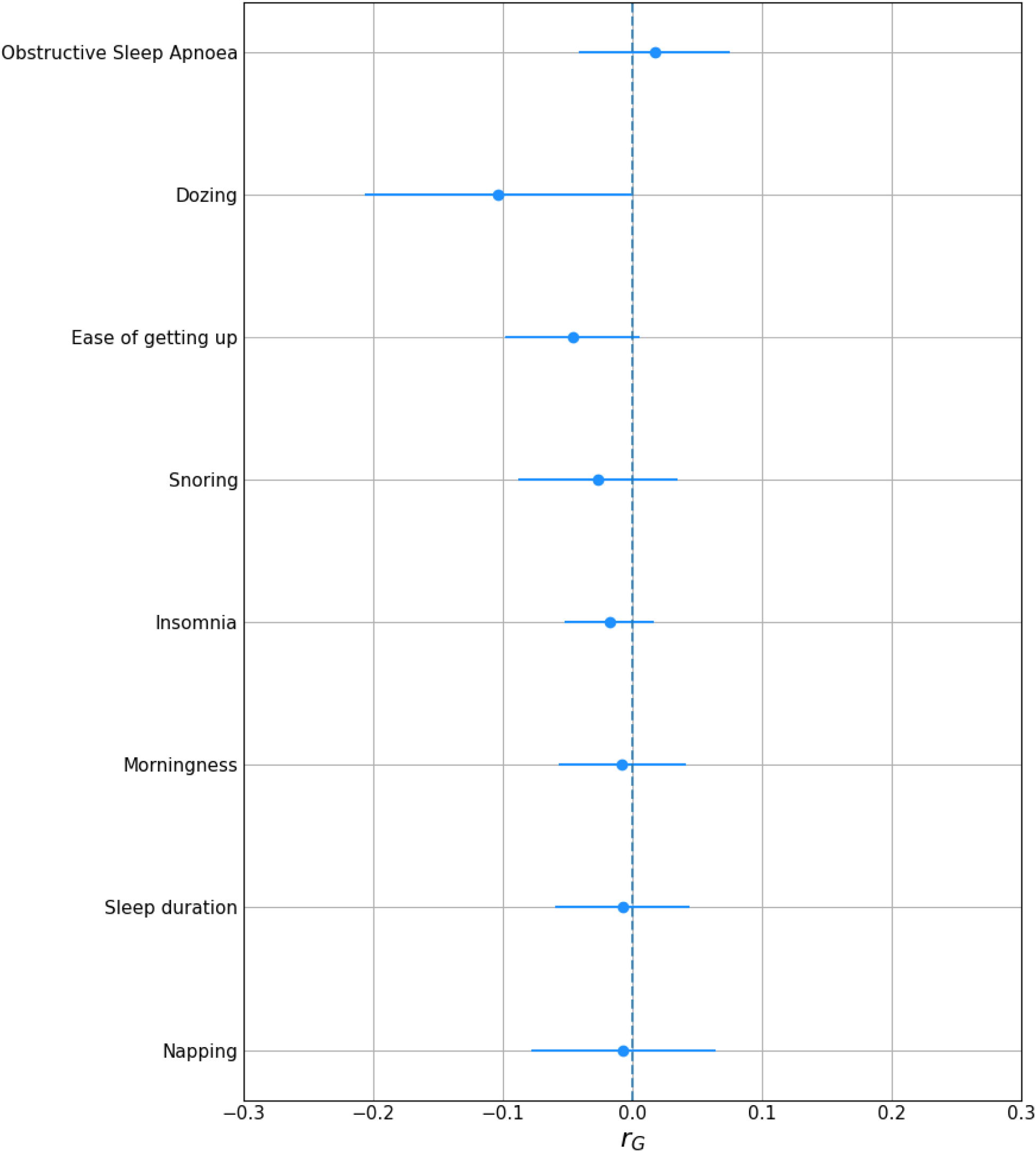
Linkage Disequilibrium Score Regression (LDSC) genetic correlation values (rG) between Parkinson’s disease and eight sleep-related traits with 95% confidence intervals.

To further investigate the shared genetic architecture at a local level, we performed GWAS-PW analyses (Supplementary Table 2). We identified six genomic segments that harboured shared causal variants between PD and at least one sleep-related trait (Figure 2). Using MAGMA, we mapped genetic variants within these segments to 25 protein-coding genes (Supplementary Tables 3 and 4). Among the sleep-related traits, ease of getting up had the highest number of shared genomic segments with PD (four segments), followed by morningness and sleep duration (two segments each) (Table 1).

**Fig 2.**
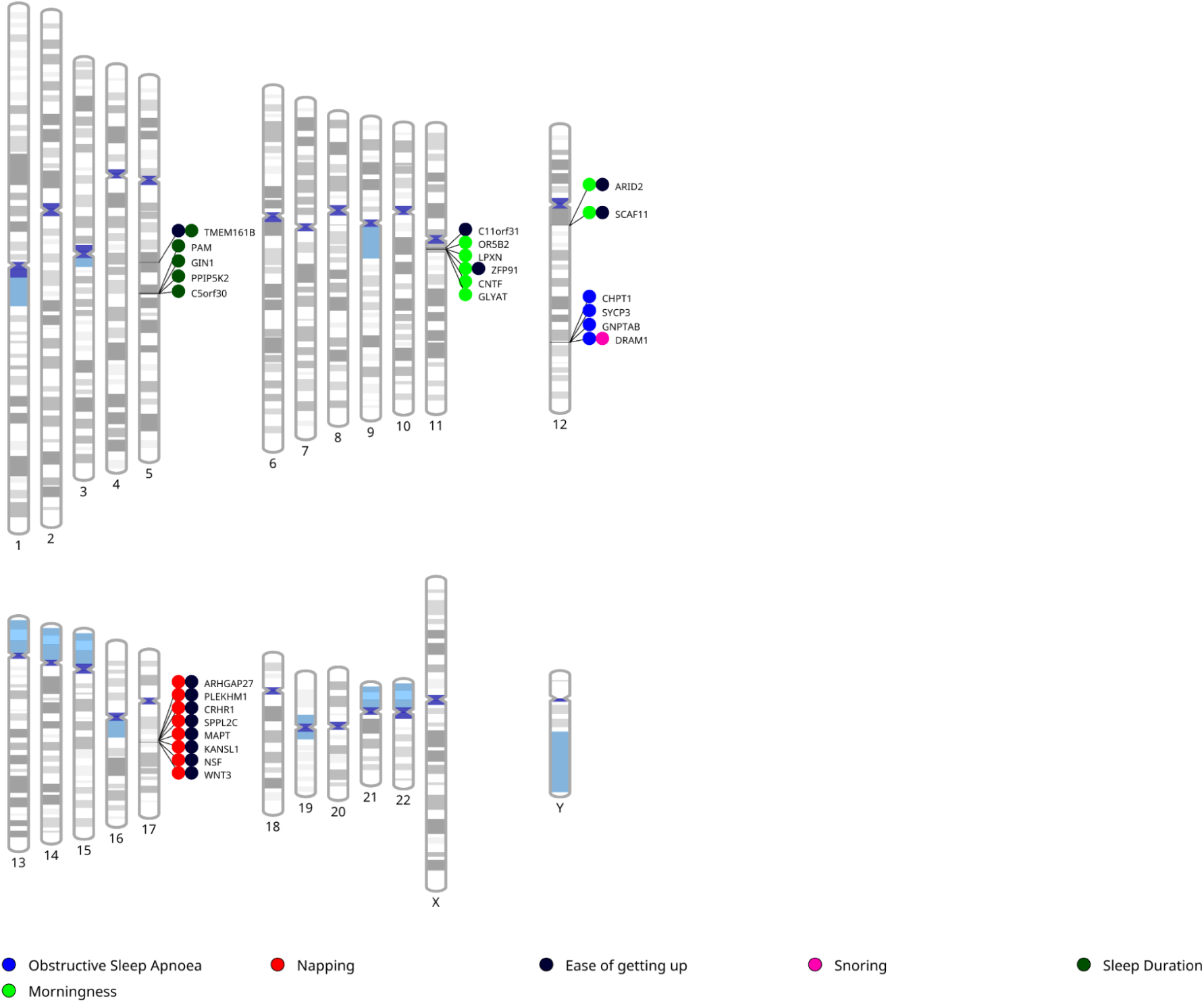
Phenogram representing genomic segments and genes shared between sleep-related traits and Parkinson’s Disease.

**Table 1.** Genomic segments with shared causal variants between Parkinson’s Disease (PD) and sleep-related traits.

| Sleep-related trait | Shared genomic segments with PD | Shared genes with PD |
| --- | --- | --- |
| Obstructive Sleep Apnoea | 1 | 4 |
| Ease of getting up | 4 | 13 |
| Morningness | 2 | 7 |
| Napping | 1 | 8 |
| Sleep duration | 2 | 5 |
| Snoring | 1 | 1 |

Multiple genes located on chromosome 17—*WNT3, CRHR1, KANSL1,* and *MAPT*—were among the most significantly associated with PD, ease of getting up, and napping. Shared loci between PD and ease of getting up were also observed on chromosomes 5, 11, and 12, whereas loci shared between PD and napping were restricted to chromosome 17. All loci associated with both PD and sleep duration were located on chromosome 5. Notably, chromosomes 5, 12, and 17 contained the top genes associated with PD (Table 2).

**Table 2.**
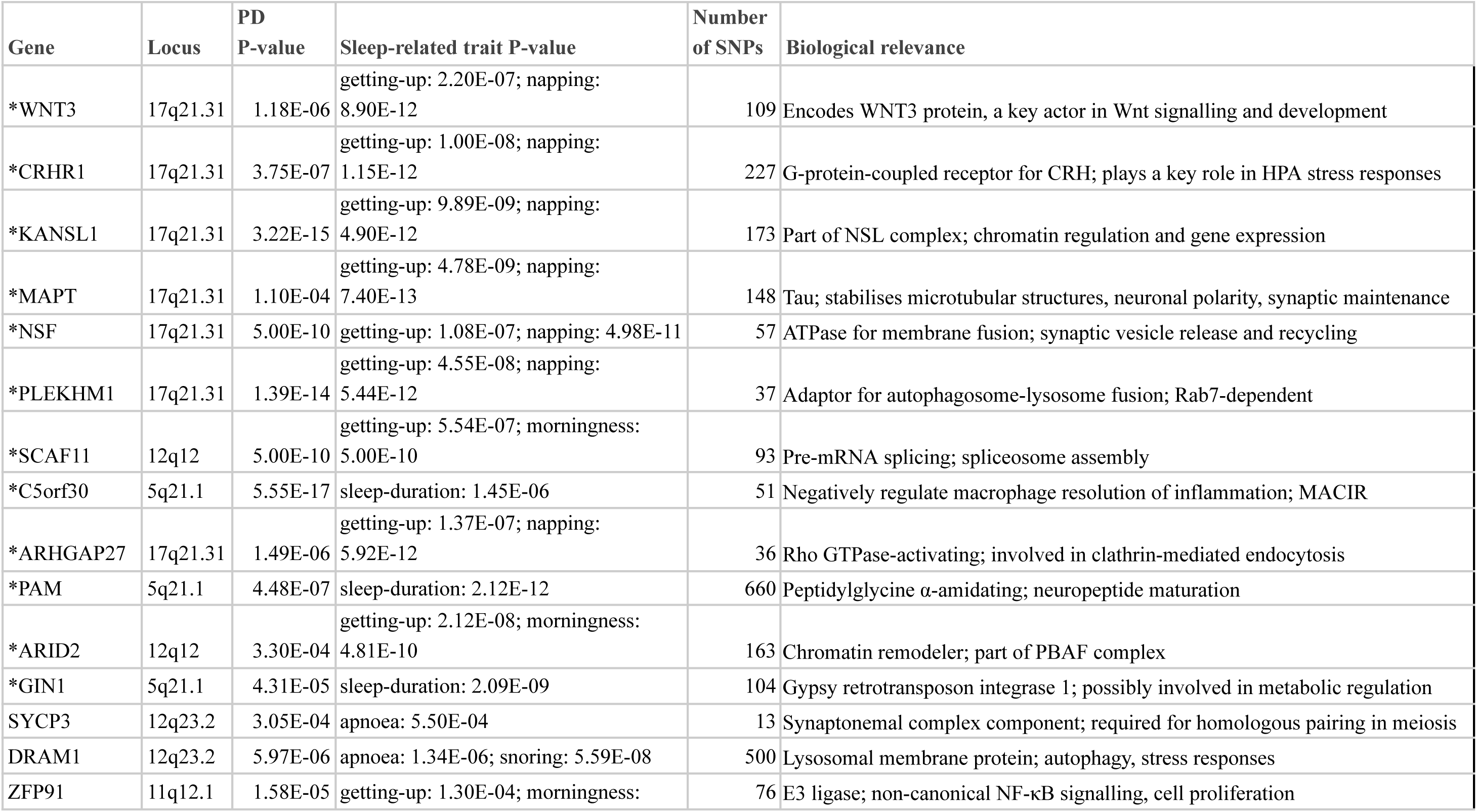
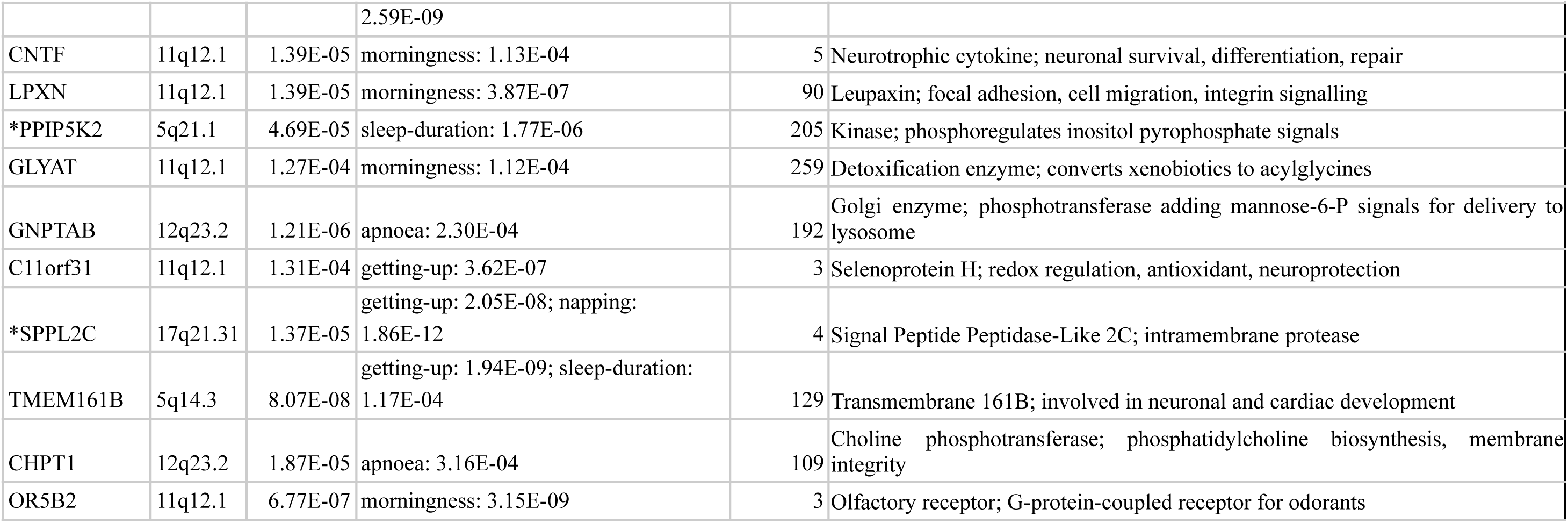
Biological relevance of the protein-coding genes associated with Parkinson’s Disease (PD) and at least one sleep-related trait. For each gene, the chromosomal locus, the gene-based P-value for PD, the associated sleep-related traits, their corresponding P-values, and the number of contributing SNPs are listed. Genes marked with an asterisk (*) are top-ranked for PD based on gene-based test significance.

We next performed gene-set enrichment analysis using MAGMA for PD and the sleep-related traits that showed the greatest number of shared genomic segments with PD: ease of getting up, napping, and sleep duration. For PD, significant enrichment was observed for pathways involved in sphingolipid metabolism, microglial proliferation, miRNA regulation, and the PI3K-AKT signalling pathway (Supplementary Table 5). For *ease of getting up*, enriched gene-sets included those related to transcriptional regulation, circadian rhythms, synaptic signalling, and neurotransmission (Supplementary Table 6). In the case of *napping*, we identified enriched pathways involved in dopaminergic neurotransmission, sleep-wake cycle regulation, neuronal lipid metabolism, intracellular signalling cascades, and inflammatory and immune-related processes (Supplementary Table 7). For *sleep duration*, enriched gene-sets were related to neuronal function, synaptic plasticity, neurotransmission, and signal processing (Supplementary Table 8).

The PPI network generated using STRING for genes mapped to the 17q21.31 locus indicated modules enriched for traits related to brain structure, mood and affective symptoms, and sleep-related traits (Supplementary Table 9). Genes such as *MAPT*, *CRHR1*, and *KANSL1* formed high-confidence interactions within the network (25 nodes, 20 edges; expected = 1; PPI enrichment p < 1.0e−16), suggesting greater connectivity than expected by chance (Figure 3). Using the MCL approach, we identified four distinct clusters. The largest cluster included genes from the 17q21.31 region, *ARHGAP27*, *CRHR1*, *MAPT*, *KANSL1*, *PLEKHM1*, *SPPL2C*, and *WNT3*, while the remaining clusters grouped genes, suggesting a shared functional role or a similar pattern of connections in the protein–protein interaction map.

**Fig 3.**
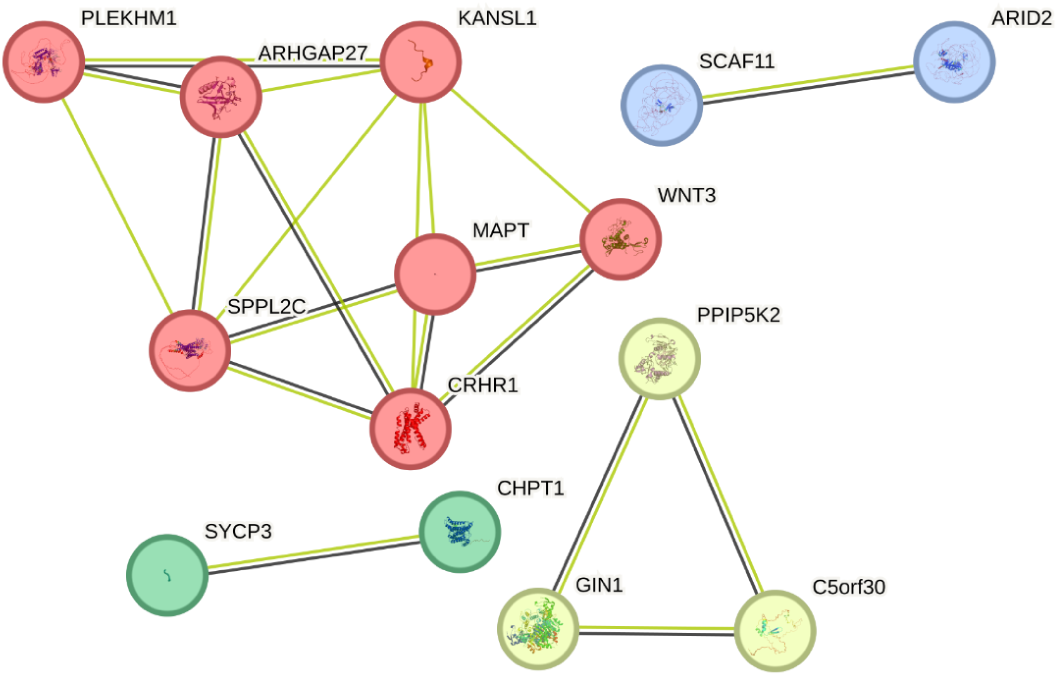
Protein-protein interaction (PPI) network of genes mapped to the 17q21.31 locus (red nodes), generated using STRING. Edges in black represent co-expression evidence, while green edges indicate associations supported by text mining. Only medium and high-confidence interactions (score ≥ 0.4) are shown.

## DISCUSSION

We investigated the shared genetic architecture between Parkinson’s disease (PD) and self-reported sleep-related traits by integrating GWAS summary statistics, conducting genetic correlation analyses, and mapping common causal variants to protein-coding genes. Although we did not detect statistically significant genome-wide genetic correlations between PD and most sleep-related traits after correcting for multiple comparisons, the GWAS-PW approach identified six genomic segments with evidence of shared causal variants. These segments were located on chromosomes 5, 11, 12, and, most prominently, chromosome 17, where the 17q21.31 locus contained the strongest associations.

Our results suggest that the relationship between PD and sleep disturbances may be mediated through specific biological axes. First, genes involved in synaptic structure and neurotransmission, such as *MAPT*, *NSF*, and *ARHGAP27*, play roles in axonal transport, vesicle trafficking, and synaptic regulation. Second, genes related to circadian and neuroendocrine regulation, *CRHR1*, *ARHGAP27*, and *WNT3*, are implicated in sleep timing, hypothalamic–pituitary–adrenal (HPA) axis activation, and circadian gene expression. Third, autophagy- and lysosomal-related genes, such as *PLEKHM1,* are key to neuronal homeostasis. Notably, *ARHGAP27* and *CRHR1* are involved in both neurotransmission and circadian regulation, suggesting that convergence across these biological domains may play an important role in linking PD to sleep-related phenotypes.

The 17q21.31 locus has previously been associated with PD risk ^30,31^. In our study, we identified multiple genes within this region—including *CRHR1, MAPT, KANSL1, NSF, WNT3, PLEKHM1*, and *ARHGAP27*—that were significantly associated with both PD and sleep traits such as ease of getting up and napping. These genes have established roles in neurodegenerative and brain-related processes, including tau pathology, synaptic transmission, stress response, and circadian regulation^32^. Additionally, their high degree of connectivity in the PPI network suggests coordinated regulation and shared participation in functional pathways relevant to both sleep and neurodegeneration.

*MAPT* and *KANSL1*, located within the 17q21.31 inversion polymorphism, have been co-implicated in PD risk via sub-haplotypes that modulate gene expression ^31^. Genes prioritised in our study, including *MAPT, KANSL1, NSF*, and *ARHGAP27*, are central to pathways supporting synaptic integrity. For example, *MAPT* encodes the tau protein, crucial for maintaining axonal structure and transport. *NSF* is essential for vesicle fusion and neurotransmitter release, while *ARHGAP27*, a Rho GTPase-activating protein, regulates cytoskeletal dynamics and neuronal signalling and has been associated with chronotype and daytime sleepiness ^33^.

*CRHR1* encodes the corticotropin-releasing hormone receptor 1, a central regulator of the HPA axis and circadian rhythms ^34,35^. Its expression has been linked to sleep timing, duration, mood regulation, and stress reactivity ^36^. Chronic HPA axis hyperactivity has been associated with both PD progression and sleep disturbances, positioning CRHR1 as a potential mechanistic link between these phenotypes ^37^. Similarly, WNT3, a component of the Wnt signalling pathway, is involved in neurodevelopment and synaptic plasticity and may also be relevant to sleep–brain interactions ^38^.

*PLEKHM1*, implicated in lysosomal trafficking and autophagy, also emerged as a candidate gene of interest. Autophagy dysfunction is a recognised hallmark of PD and has been linked to disrupted sleep regulation ^39,40^. Impairment in lysosomal clearance may lead to alpha-synuclein accumulation and neuroinflammation, key mechanisms in PD pathogenesis that are also modulated by sleep^41^. Some of the enriched pathways in our study, including PI3K-AKT signalling, sphingolipid metabolism, and circadian regulation, have also been shown to involve astrocytic activity ^42–44^.

Gene-set enrichment analyses revealed convergence on pathways involved in synaptic and neurotransmission functions across PD, ease of getting up, and sleep duration, highlighting genes such as *NSF*, *MAPT*, and *ARHGAP27*. In addition, gene sets associated with dopaminergic and adrenergic signalling were significantly enriched in comparisons involving sleep duration. These findings suggest that shared genetic mechanisms may involve both neurotransmission and circadian processes.

We also identified enrichment of a gene set containing downregulated targets of microRNA miR-1 in both PD and sleep duration, suggesting a possible role for miRNA-mediated post-transcriptional regulation. While miR-1 is best known for its role in muscle physiology, emerging evidence suggests it may also influence neuronal development and circadian control ^45,46^.

Several gene sets were shared across ease of getting up, napping, and sleep duration. For example, pathways related to sleep regulation were enriched for both ease of getting up and napping, consistent with gene-level findings implicating *CRHR1* in circadian and neuroendocrine regulation. The overlap of pathways involved in circadian regulation, synaptic function, and HPA axis activity suggests a biological basis for the relationship between sleep traits and vulnerability to neurodegeneration. These results align with the hypothesis of a bidirectional association between PD and sleep disturbances^47^, in which shared genetic factors contribute to both neurodegenerative risk and susceptibility to sleep dysregulation.

There are several limitations to our study. First, the GWAS summary statistics were restricted to individuals of European ancestry, which limits the generalisability of our findings to other populations. Second, the 17q21.31 region is characterised by a large inversion polymorphism and extensive linkage disequilibrium, complicating efforts to fine-map causal variants and their regulatory targets. Third, while gene-set enrichment analysis provides biological context, its interpretability is constrained by current limitations in gene-set annotation, and some pathways may be incompletely represented ^48^.

In summary, we provide evidence for shared genetic loci between Parkinson’s disease and multiple sleep-related traits, particularly within the 17q21.31 region. This locus contains several genes with roles in neurodegeneration, circadian regulation, and stress response. Our findings support the hypothesis that sleep disturbances in PD may reflect not only disease progression but also shared aetiological pathways. Converging results from gene-set analyses suggest that neurotransmission, circadian regulation, and neuroinflammatory processes are potential mechanisms underlying this overlap. These findings underscore the relevance of sleep-related endophenotypes in the genetic architecture of PD, suggesting new directions for early detection and mechanistic studies.

## Author Contributions

All authors contributed significantly to the work reported, whether in the conception, study design, execution, acquisition of data, analysis and interpretation, or all these areas; took part in drafting, revising or critically reviewing the article; gave final approval of the version to be published; and have agreed on the journal to which the article has been submitted.

## Data Availability

The full GWAS summary statistics for PD and sleep-related traits are available upon request through a Data Transfer Agreement and application procedure, including the 23andMe cohort (https://research.23andme.com/dataset-access/).

## Code Availability

No custom code was implemented in this study. All genetic analyses were performed using publicly available software, which is cited and described in the main text.

## Ethics declaration

This study was conducted under the oversight of QIMR Berghofer’s Human Research Ethics Committee. All participants provided informed consent.

UK Biobank: The UK Biobank study was approved by the National Health Service National Research Ethics Service (ref. 11/NW/0382), and all participants gave their written consent to take part in the UK Biobank study. Further details about ethics oversight in UK Biobank can be found at https://www.ukbiobank.ac.uk/ethics/.

23andMe Inc: Participants provided consent and took part in the research online under a protocol approved by the independent AAHRPP-accredited Institutional Review Board, Ethical & Independent Review Services (E&I Review).

## Competing Interests

All authors declare no conflicts of interest.

## Notes

### Competing Interest Statement

The authors have declared no competing interest.

### Funding Statement

This study did not receive any funding

### Author Declarations

The Human Research Ethics Committee of the Queensland Institute of Medical Research Berghofer gave ethical approval for this work.

## REFERENCES

1. Pringsheim, T., Jette, N., Frolkis, A. & Steeves, T. D. L. The prevalence of Parkinson’s disease: a systematic review and meta-analysis: PD PREVALENCE. Mov. Disord. 29, 1583–1590 (2014).

2. Váradi, C. Clinical features of Parkinson’s disease: The evolution of critical symptoms. Biology (Basel) 9, 103 (2020).

3. Stefani, A. & Högl, B. Sleep in Parkinson’s disease. Neuropsychopharmacology 45, 121–128 (2020).

4. Naia, C. V. et al. Sleep disturbances in Parkinson’s disease: A scoping review. Braz. J. Clin. Med. Rev. 3, bjcmr22 (2025).

5. Dhawan, V., Healy, D. G., Pal, S. & Chaudhuri, K. R. Sleep-related problems of Parkinson’s disease. Age Ageing 35, 220–228 (2006).

6. Iranzo, A., Cochen De Cock, V., Fantini, M. L., Pérez-Carbonell, L. & Trotti, L. M. Sleep and sleep disorders in people with Parkinson’s disease. Lancet Neurol. 23, 925–937 (2024).

7. Barber, A. & Dashtipour, K. Sleep disturbances in Parkinson’s disease with emphasis on rapid eye movement sleep behavior disorder. Int. J. Neurosci. 122, 407–412 (2012).

8. Bohnen, N. I. & Hu, M. T. M. Sleep disturbance as potential risk and progression factor for Parkinson’s disease. J. Parkinsons. Dis. 9, 603–614 (2019).

9. Lysen, T. S., Darweesh, S. K. L., Ikram, M. K., Luik, A. I. & Ikram, M. A. Sleep and risk of parkinsonism and Parkinson’s disease: a population-based study. Brain 142, 2013–2022 (2019).

10. Kocevska, D., Barclay, N. L., Bramer, W. M., Gehrman, P. R. & Van Someren, E. J. W. Heritability of sleep duration and quality: A systematic review and meta-analysis. Sleep Med. Rev. 59, 101448 (2021).

11. Lind, M. J., Aggen, S. H., Kirkpatrick, R. M., Kendler, K. S. & Amstadter, A. B. A longitudinal twin study of insomnia symptoms in adults. Sleep 38, 1423–1430 (2015).

12. Lopez-Minguez, J., Morosoli, J. J., Madrid, J. A., Garaulet, M. & Ordoñana, J. R. Heritability of siesta and night-time sleep as continuously assessed by a circadian-related integrated measure. Sci. Rep. 7, 12340 (2017).

13. Blauwendraat, C., Nalls, M. A. & Singleton, A. B. The genetic architecture of Parkinson’s disease. Lancet Neurol. 19, 170–178 (2020).

14. Nalls, M. A. et al. Identification of novel risk loci, causal insights, and heritable risk for Parkinson’s disease: a meta-analysis of genome-wide association studies. Lancet Neurol. 18, 1091–1102 (2019).

15. Jansen, P. R. et al. Genome-wide analysis of insomnia in 1,331,010 individuals identifies new risk loci and functional pathways. Nat. Genet. 51, 394–403 (2019).

16. Watanabe, K. et al. Genome-wide meta-analysis of insomnia prioritizes genes associated with metabolic and psychiatric pathways. Nat. Genet. 54, 1125–1132 (2022).

17. Willer, C. J., Li, Y. & Abecasis, G. R. METAL: fast and efficient meta-analysis of genomewide association scans. Bioinformatics 26, 2190–2191 (2010).

18. Campos, A. I. et al. Discovery of genomic loci associated with sleep apnea risk through multi-trait GWAS analysis with snoring. Sleep 46, zsac308 (2023).

19. Campos, A. I. et al. Insights into the aetiology of snoring from observational and genetic investigations in the UK Biobank. Nat. Commun. 11, 817 (2020).

20. Bulik-Sullivan, B. K. et al. LD Score regression distinguishes confounding from polygenicity in genome-wide association studies. Nat. Genet. 47, 291–295 (2015).

21. Pickrell, J. K. et al. Detection and interpretation of shared genetic influences on 42 human traits. Nat. Genet. 48, 709–717 (2016).

22. García-Marín, L. M. et al. Phenome-wide screening of GWAS data reveals the complex causal architecture of obesity. Hum. Genet. 140, 1253–1265 (2021).

23. Aman, A. M. et al. Phenome-wide screening of the putative causal determinants of depression using genetic data. Hum. Mol. Genet. 31, 2887–2898 (2022).

24. García-Marín, L. M. et al. Large-scale genetic investigation reveals genetic liability to multiple complex traits influencing a higher risk of ADHD. Sci. Rep. 11, 22628 (2021).

25. García-Marín, L. M., Campos, A. I., Martin, N. G., Cuéllar-Partida, G. & Rentería, M. E. Phenome-wide analysis highlights putative causal relationships between self-reported migraine and other complex traits. J. Headache Pain 22, 66 (2021).

26. de Leeuw, C. A., Mooij, J. M., Heskes, T. & Posthuma, D. MAGMA: generalized gene-set analysis of GWAS data. PLoS Comput. Biol. 11, e1004219 (2015).

27. Watanabe, K., Taskesen, E., van Bochoven, A. & Posthuma, D. Functional mapping and annotation of genetic associations with FUMA. Nat. Commun. 8, 1826 (2017).

28. Reyes-Pérez, P. et al. Investigating the shared genetic etiology between Parkinson’s disease and depression. J. Parkinsons. Dis. 14, 483–493 (2024).

29. Szklarczyk, D. et al. The STRING database in 2023: protein-protein association networks and functional enrichment analyses for any sequenced genome of interest. Nucleic Acids Res. 51, D638–D646 (2023).

30. Harerimana, N. V., Goate, A. M. & Bowles, K. R. The influence of 17q21.31 and APOE genetic ancestry on neurodegenerative disease risk. Front. Aging Neurosci. 14, 1021918 (2022).

31. Bowles, K. R. et al. 17q21.31 sub-haplotypes underlying H1-associated risk for Parkinson’s disease are associated with LRRC37A/2 expression in astrocytes. Mol. Neurodegener. 17, 48 (2022).

32. Koopman-Verhoeff, M. E. et al. Genome-wide DNA methylation patterns associated with sleep and mental health in children: a population-based study. J. Child Psychol. Psychiatry 61, 1061–1069 (2020).

33. Moon, S. Y. & Zheng, Y. Rho GTPase-activating proteins in cell regulation. Trends Cell Biol. 13, 13–22 (2003).

34. Diaz-Torres, S. et al. Macular structural integrity estimates are associated with Parkinson’s disease genetic risk. Acta Neuropathol. Commun. 12, 130 (2024).

35. Yang, Y. et al. Chronic corticosterone disrupts the circadian rhythm of CRH expression and m6A RNA methylation in the chicken hypothalamus. J. Anim. Sci. Biotechnol. 13, 29 (2022).

36. Wang, T. et al. The nucleus accumbens CRH-CRHR1 system mediates early-life stress-induced sleep disturbance and dendritic atrophy in the adult mouse. Neurosci. Bull. 39, 41–56 (2023).

37. García-Marín, L. M. et al. Shared molecular genetic factors influence subcortical brain morphometry and Parkinson’s disease risk. NPJ Parkinsons Dis. 9, 73 (2023).

38. Maguschak, K. A. & Ressler, K. J. A role for WNT/β-catenin signaling in the neural mechanisms of behavior. J. Neuroimmune Pharmacol. 7, 763–773 (2012).

39. Lu, J., Wu, M. & Yue, Z. Autophagy and Parkinson’s disease. Adv. Exp. Med. Biol. 1207, 21–51 (2020).

40. Chauhan, A. K. & Mallick, B. N. Association between autophagy and rapid eye movement sleep loss-associated neurodegenerative and patho-physio-behavioral changes. Sleep Med. 63, 29–37 (2019).

41. Pajares, M., I Rojo, A., Manda, G., Boscá, L. & Cuadrado, A. Inflammation in Parkinson’s disease: Mechanisms and therapeutic implications. Cells 9, 1687 (2020).

42. Sun, Y.-Y. et al. Cell type-specific dependency on the PI3K/Akt signaling pathway for the endogenous Epo and VEGF induction by baicalein in neurons versus astrocytes. PLoS One 8, e69019 (2013).

43. Alaamery, M. et al. Role of sphingolipid metabolism in neurodegeneration. J. Neurochem. 158, 25–35 (2021).

44. Carvalhas-Almeida, C. & Sehgal, A. Glia: the cellular glue that binds circadian rhythms and sleep. Sleep 48, zsae314 (2025).

45. Safa, A. et al. miR-1: A comprehensive review of its role in normal development and diverse disorders. Biomed. Pharmacother. 132, 110903 (2020).

46. Na, Y. J. et al. Comprehensive analysis of microRNA-mRNA co-expression in circadian rhythm. Exp. Mol. Med. 41, 638–647 (2009).

47. Minakawa, E. N. Bidirectional relationship between sleep disturbances and Parkinson’s disease. Front. Neurol. 13, 927994 (2022).

48. Mubeen, S., Tom Kodamullil, A., Hofmann-Apitius, M. & Domingo-Fernández, D. On the influence of several factors on pathway enrichment analysis. Brief. Bioinform. 23, bbac143 (2022).

